# Prioritizing Parkinson’s disease risk genes in genome-wide association loci

**DOI:** 10.1101/2024.12.13.24318996

**Authors:** Lara M. Lange, Catalina Cerquera-Cleves, Marijn Schipper, Georgia Panagiotaropoulou, Alice Braun, Julia Kraft, Swapnil Awasthi, Nathaniel Bell, Danielle Posthuma, Stephan Ripke, Cornelis Blauwendraat, Karl Heilbron

## Abstract

Recent advancements in Parkinson’s disease (PD) drug development have been significantly driven by genetic research. Importantly, drugs supported by genetic evidence are more likely to be approved. While genome-wide association studies (GWAS) are a powerful tool to nominate genomic regions associated with certain traits or diseases, pinpointing the causal biologically relevant gene is often challenging. Our aim was to prioritize genes underlying PD GWAS signals.

The polygenic priority score (PoPS) is a similarity-based gene prioritization method that integrates genome-wide information from MAGMA gene-level association tests and more than 57,000 gene-level features, including gene expression, biological pathways, and protein-protein interactions. We applied PoPS to data from the largest published PD GWAS in East Asian- and European-ancestries.

We identified 120 independent associations with *P* < 5×10^−8^ and prioritized 46 PD genes across these loci based on their PoPS scores, distance to the GWAS signal, and presence of non-synonymous variants in the credible set. Alongside well-established PD genes (*e.g., TMEM175* and *VPS13C*), some of which are targeted in ongoing clinical trials (*i.e.*, *SNCA*, *LRRK2*, and *GBA1*), we prioritized genes with a plausible mechanistic link to PD pathogenesis (*e.g., RIT2, BAG3*, and *SCARB2*). Many of these genes hold potential for drug repurposing or novel therapeutic developments for PD (*i.e., FYN, DYRK1A, NOD2, CTSB, SV2C,* and *ITPKB*). Additionally, we prioritized potentially druggable genes that are relatively unexplored in PD (*XPO1, PIK3CA, EP300, MAP4K4, CAMK2D, NCOR1,* and *WDR43*).

We prioritized a high-confidence list of genes with strong links to PD pathogenesis that may represent our next-best candidates for disease-modifying therapeutics. We hope our findings stimulate further investigations and preclinical work to facilitate PD drug development programs.

## Introduction

Parkinson’s disease (PD) is a neurodegenerative condition with the fastest-growing global prevalence among all neurological disorders, posing significant challenges to healthcare systems and society^1^. The pathophysiology of PD involves multiple risk factors, including genetics, environmental factors, and aging^2^. Currently, most therapeutic strategies aim to address dopamine deficiency (*e.g.,* levodopa or dopamine agonists). More advanced therapies, such as deep brain stimulation, focus on modulating neural synchrony in distributed brain networks^3^. Although these treatments are intended to alleviate symptoms, they do not clearly impact disease progression^4^. Hence, there is an urgent need to identify specific therapeutic targets that can effectively halt the course of the disease and accelerate the clinical development of disease-modifying therapies.

Recent advancements in drug development for PD have been significantly influenced by over two decades of genetic research. This body of evidence has not only deepened our understanding of the biology underlying PD, but has also led to the identification of numerous drug targets. Some of these are currently being evaluated in clinical trials^5^, including *GBA1*, *LRRK2* and *SNCA*. Drugs supported by genetic evidence are more than twice as likely to be approved^6,7^, and human genetic data supported two-thirds of the drugs approved by the FDA from 2013 to 2022^8,9^. Leveraging data from genetic studies, and developing pipelines to prioritize and select the best targets is estimated to significantly impact the development of new drugs^10^. This is especially true for disease-modifying therapies^6,11^.

In recent years, various genome-wide association studies (GWAS) have identified over 100 loci associated with PD^12–18^. The largest European-ancestry PD GWAS identified 90 independent significant risk signals across 78 genomic regions (37,688 cases, 18,618 first-degree relatives of cases, and 1,417,791 controls)^13^. A polygenic risk score including 1,805 variants explained 16-36% of the heritable risk of PD^13^. Additionally, 15 population-specific variants were identified through GWAS conducted in individuals of Latino, East Asian, South Asian, and African and African admixed ancestry^14–17^. A recent large-scale multi-ancestry meta-analysis of PD GWAS uncovered 12 potentially novel loci and fine-mapped six putative causal variants at six previously identified loci (49,049 cases and 2,452,961 controls from these four ancestral populations)^18^.

A major limitation of GWAS is that it nominates genomic regions, not the specific genes that confer risk. Typically, GWAS hits can be explained by two general mechanisms: 1) a functional coding variant that alters protein function or 2) a non-coding variant that regulates transcription or translation. Often GWAS studies annotate their loci with the closest gene to the lead variant. This is the correct causal gene strikingly often^19^, although more sophisticated methods have been developed to improve predictive accuracy.

There have been previous gene prioritization efforts using machine learning models that were trained on and applied to non-PD traits (“locus-to-gene” (L2G) model ^20^) or PD specifically (referred to as Yu2024^21^). However, all previous efforts to prioritize PD GWAS genes have done so on a locus-by-locus basis, ignoring information from other significant loci and the rest of the genome. The polygenic priority score (PoPS)^19^ is a gene prioritization tool that incorporates genome-wide information from MAGMA^22^ gene-level association tests and more than 57,000 gene-level features (*i.e.,* gene expression, biological pathways, and protein-protein interactions). By intersecting genes with the top PoPS value in their locus with genes that were nearest to their GWAS signal, they were able to achieve a precision of 79%.

Here we applied PoPS to several PD GWAS summary statistics including European and East Asian ancestries and highlight high-confidence prioritized genes.

## Methods and Materials

### Ethics statement

This study was conducted in accordance with the ethical standards of the institutional and national research committees. Details on Institutional Review Board approvals of the individual studies included in the presented work are provided in the original publications^13,16^.

### GWAS summary statistics

We analyzed two published GWAS datasets. First, an East Asian-ancestry (EAS) meta-analysis of 6,724 cases and 24,851 controls (effective sample size [*N*_eff_] = 15,886)^16^. Second, a European-ancestry (EUR) meta-analysis of 37,688 cases, 18,618 proxy cases (first degree relative with PD), and 1,417,791 controls (*N*_eff_ = 127,626)^13^. We performed a fixed-effects meta-analysis of these two datasets using METAL^23^ to generate a combined EAS+EUR dataset.

### Reference panels

We used all available data from the Haplotype Reference Consortium release 1.1 (HRC) to construct three linkage disequilibrium (LD) reference panels: an EAS panel (*N* = 538), an EUR panel (*N* = 16,860), and an EAS+EUR panel that included both EAS and EUR individuals in the same proportions as the GWAS summary statistics - 11% EAS and 89% EUR (*N*_EUR_ = 4,322, *N*_EAS_ = 538).

### Variant quality control

We removed EUR GWAS variants with: 1) a reported allele frequency that differed from the EUR reference panel frequency by > 0.1 (42 variants removed) and 2) a reported allele frequency that differed from the EUR reference panel frequency by > 12-fold (an additional 462 variants removed). No variants failed these same quality control checks for the EAS GWAS and EAS+EUR meta-analysis. After quality control, the number of variants remaining was 5,347,472 for EAS, 6,584,031 for EUR, and 7,593,632 for EAS+EUR.

### Isolating independent association signals

In order to disentangle statistically-independent genetic signals in the EAS+EUR dataset, we first clumped variants using PLINK v1.9^24^ (*P* < 5×10^−8^, *r*^2^ < 0.1, window size = 3 Mbp) and our EAS+EUR reference panel, expanded the boundaries of each clump by 500kb on either side, and merged overlapping boundaries. Within each resulting region, we ran COJO^25^ and removed hits with joint *P* > 5×10^−8^. If multiple independent hits in a region were found, we used COJO to isolate each signal by performing leave-one-hit-out conditional analysis. For each isolated signal, we computed credible sets (CSs) using the finemap.abf function in the coloc R package^26,27^. Finally, we defined loci as ± 300 kb around each credible set.

### MAGMA and PoPS

We performed gene-based association tests using MAGMA^22^ (“SNP-wise mean model”) and all variants with MAF > 1%. We analyzed the EAS- and EUR-based GWAS separately using the corresponding ancestry-specific reference panel and MAFs. We mapped variants to protein-coding genes using Genome Reference Consortium Human Build 37 (GRCh37) gene start and end positions from GENCODE v44^28^. We removed genes that had fewer than three variants mapped to them. For each gene, we meta-analyzed the resulting ancestry-specific MAGMA z-scores weighted by the square root of sample size^23^. Using the ancestry-specific MAGMA results as input, we performed PoPS^19^ using all 57,543 gene-based features as predictors. These features were not available for chrX so we restricted our analysis to autosomal genes. The resulting ancestry-specific PoPS values were then also meta-analyzed weighted by the square root of sample size. We only used the meta-analyzed MAGMA and PoPS values for gene prioritization.

### Gene prioritization criteria

Following the original PoPS publication, we prioritized genes that met both of the following criteria: 1) had the top PoPS value in a given locus and 2) were the nearest gene to the corresponding GWAS signal based on the posterior inclusion probability (PIP)-weighted average position of credible set variants (the smallest set of variants expected to contain the causal variant with 95% probability). We also prioritized genes that had PIP > 50% for non-synonymous credible set variants affecting the gene. We used non-synonymous variants from the “baseline-LF 2.2.UKB model” (80,693 variants)^29^. If a locus contained multiple prioritized genes, we only included the gene that was prioritized due to non-synonymous credible set variants.

### Comparison with previous PD gene prioritization efforts

We compared our prioritized genes with those highlighted in two previous studies: Mountjoy *et al.* 2021^20^ and Yu *et al.* 2024^21^. Both studies trained XGBoost models to predict the probability that a given gene is causal in a given GWAS locus, and have been applied to the EUR PD GWAS data. We downloaded results for the Mountjoy *et al.* 2021 study^20^ from the Open Targets Genetics website (https://genetics.opentargets.org/Study/GCST009325/). We extracted results for the Yu *et al.* 2024^21^ study from Supplementary Table 2. Although there only appears to be one GWAS hit near the *FCGR2A* gene in the EUR dataset^13^, Yu *et al.* 2024^21^ causal probabilities in this locus sum to over 2500%. If the sum of Yu *et al.* 2024^21^ causal probabilities within a given locus exceeded 100%, we therefore rescaled them to sum to 100%.

### Drug repurposing and tractability

We determined whether our prioritized genes were targeted by approved or investigational drugs using GraphQL API queries of the Open Targets platform^30^, which in turn queries the EMBL-EBI ChEMBL database. For genes that were not targeted by approved or investigational drugs, we performed additional Open Targets API queries to extract evidence of drug tractability—the probability of identifying a small molecule drug that is able to bind and modulate a given target. We considered genes to encode proteins that may be druggable if they have been co-crystallized with a small molecule.

### Literature review

We performed a PubMed search (https://pubmed.ncbi.nlm.nih.gov/) for each prioritized gene using the standardized search term: “GENE_NAME[Title/Abstract] AND parkinson[Title/Abstract]”. We excluded known monogenic or high-risk genes: *LRRK2* (OMIM: 607060)*, GBA1* (OMIM: 168600)*, SNCA* (OMIM: 168601), and *VPS13C* (OMIM: 616840). We also excluded *TMEM175*, which has been experimentally confirmed as the causal gene in its locus. For each search that returned at least eight publications, we performed a more detailed literature review. We excluded genes with fewer than eight publications from further analyses. We screened the abstracts for evidence of a potential role of each gene in the pathogenesis of PD and information regarding the potential mechanism of action, thereby nominating it as a potential drug target. If required, an additional full-text review was performed.

## Results

### Methods overview

We prioritized PD genes using a meta-analysis of the largest published GWASes in East Asian-^16^ and European-ancestry^13^ individuals (44,412 cases, 18,618 proxy cases, and 1,442,642 controls). We identified 120 independent associations with *P* < 5×10^−8^ (Supplementary Table 1). Across these loci, we prioritized 46 PD genes (Fig. 1, Supplementary Table 2) based on their PoPS scores, distance to the credible set, and presence of non-synonymous variants in the credible set. We further compared our findings with prioritization efforts using the L2G model^20^ and from Yu *et al.* 2024^21^ (Fig. 1).

**Figure 1.**
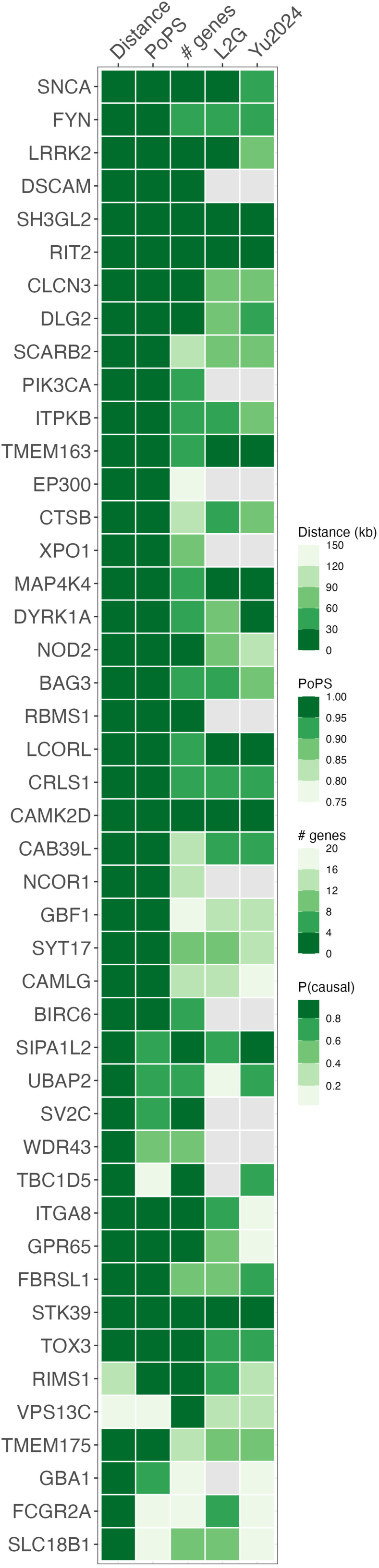
Heatmap of prioritized Parkinson’s disease genes. An overview of the evidence supporting each prioritized gene. Distance: distance in kilobases between gene and credible set. PoPS: PoPS percentile where 0 represents the smallest genome-wide value and 1 represents the largest. MAGMA: # genes: number of genes in the locus. L2G: Probability of being the causal genes according to the L2G model ^20^. Yu2024: Probability of being the causal genes according to the Yu2024 model ^21^.

### Many prioritized genes are also supported by previous literature

Our analysis prioritized several known monogenic or high-risk PD genes, including *SNCA* (OMIM: 168601), *LRRK2* (OMIM: 607060), *GBA1* (OMIM: 168600), and *VPS13C* (OMIM: 616840). For some of these, PD clinical trials are already ongoing. *LRRK2,* thought to cause PD through a gain-of-function mechanism leading to increased LRRK2 kinase activity, is being targeted by kinase inhibitors^5^. In contrast, other therapies aim to activate glucocerebrosidase (GCase), encoded by *GBA1*, and increase its activity^31,32^. Furthermore, there are different approaches of targeting α-synuclein (encoded by *SNCA*), *e.g.*, by reducing extracellular α-synuclein (PASADENA^33^ and SPARK^34,35^ trials) or blocking misfolding^36^ or aggregation^37^. We also prioritized *TMEM175*, which has been experimentally validated to be the causal gene in its respective GWAS locus^38^.

We identified several genes that were also supported by our literature review. Among these, *RIT2, DYRK1A, BAG3*, and *SCARB2* were particularly promising, with strong evidence highlighting their involvement in PD pathogenesis, although notably, no focussed locus dissection efforts have been performed yet. *RIT2* is linked to neuroprotective pathways in dopaminergic neurons^39^. Reduced RIT2 expression increases neurodegeneration in various preclinical PD models^40–43^, while RIT2 overexpression rescues autophagy-lysosome deficits and reduces α-synuclein aggregation^40,42^. *DYRK1A* has been linked to several neurodegeneration pathways^44,45^. DYRK1A appears to regulate α-synuclein inclusion formation^46^ and directly phosphorylate (and thereby inhibit) parkin^47^. *BAG3* plays a crucial role in macroautophagy and the clearance of alpha-synuclein aggregates^48,49^. *BAG3* overexpression also suppresses neuroinflammation, decreasing the release of pro-inflammatory cytokines and contributing to mitigating neuronal damage in PD^50–52^. *SCARB2* is essential for trafficking GCase from the endoplasmic reticulum to lysosomes^53^. Deficiencies in *SCARB2* lead to reduced GCase activity and subsequent α-synuclein accumulation, contributing to neurotoxicity in dopaminergic neurons^53–57^. Biallelic variants in *SCARB2* cause progressive myoclonic epilepsy with or without renal failure.

Furthermore, we prioritized several genes without strong literature evidence but predicted to be the most likely gene responsible underlying the PD GWAS signal with a probability greater than 80% by L2G or Yu2024 (*SIPA1L2, SH3GL2*, *TMEM163*, *MAP4K4*, *LCORL*, *CAMK2D*, *STK39*)^20,21^ (Fig. 1).

### Prioritized genes that encode potentially druggable targets for PD

Of the 46 prioritized genes, six (*FYN, DYRK1A, NOD2, CTSB, SV2C, ITPKB)* were identified as promising drug targets for PD, each supported by at least eight PD-related publications in our literature review. Their potential as therapeutic targets was established based either on their suitability for repurposing existing drugs to target them in PD (Supplementary Table 3) or their promise for new drug development using the Open Targets platform.

FYN inhibitors have been approved to treat various cancers (e.g., chronic myeloid leukemia and acute lymphoblastic leukemia)^58,59^ and tested in clinical trials for Alzheimer disease^60–62^. *FYN* plays a significant role in neuroinflammation and protein aggregation^63–65^. Inhibition of FYN has shown promise in reducing these detrimental processes and alleviating L-dopa-induced dyskinesia by regulating the phosphorylation of the NMDA receptor^66–68^.

*DYRK1A* encodes a dual-specificity kinase with a wide range of functions and interaction partners^45^. Although no approved drug targeting *DYRK1A* exists for PD, a number of DYRK1A inhibitors have been investigated in the context of other neurological diseases^69–72^. DYRK1A seems to directly phosphorylate a-synuclein and thereby affect aggregate formation and cell death in immortalized hippocampal neurons and brain tissue samples from a MPTP-induced PD mouse model, whereas DYRK1A inhibition suppressed a-synuclein aggregation and reduced dopaminergic neuron apoptosis^46,73^.

*CTSB* encodes cathepsin B (catB), a lysosomal enzyme crucial for protein degradation and regulating autophagy. CatB knock-out or inhibition impairs lysosomal functions in dopaminergic neurons, leads to reduced GCase activity^74,75^, and promotes α-synuclein aggregation^74^. However, farnesyltransferase inhibitors enhance cathepsin transport and activity, thereby improving lysosomal function and α-synuclein clearance^76^. These inhibitors, already approved for Hutchinson-Gilford progeria syndrome^77^, could have potential for repurposing in PD.

Although there is an approved drug targeting *NOD2* for osteosarcoma^78^, it acts as an activator and induces a proinflammatory response. However, data suggest that *NOD2* inhibition and an anti-inflammatory response would be desired in PD. In an experimental mouse PD model induced by the neurotoxin 6-hydroxydopamine, NOD2 deficiency was associated with an attenuated inflammatory response and suggested to have protective effects against degeneration of dopaminergic neurons and neuronal death^79^.

*SV2C* encodes synaptic vesicle glycoprotein 2C and is involved in synaptic vesicular function. *SV2C* expression is enriched in the basal ganglia, particularly in dopaminergic neurons in the substantia nigra^80^, and is believed to be crucial for dopamine neuron function and homeostasis^81,82^. In a mouse model, *SV2C* knock-out resulted in reduced dopamine release in the dorsal striatum, disrupted α-synuclein expression, and mild motor deficits^82^, suggesting that enhancing *SV2C* function is beneficial for PD. Clinical trials targeting *SV2C* are currently ongoing for epilepsy, although it is unclear whether the drug is an activator or inhibitor. Should further preclinical testing find that it is an activator, this drug could potentially be considered for repurposing for PD.

*ITPKB* encodes a kinase involved in calcium homeostasis^83^. While it is often possible to inhibit kinases, inhibiting ITPKB is predicted to be detrimental in PD since it has been associated with increased α-synuclein aggregation and impaired autophagy^84^. However, previous work has shown that *MICU3* (a brain-specific mitochondrial calcium uniporter) operates downstream of ITPKB and inhibiting MICU3 was able to rescue increased ITPKB activity^85^. *MICU3*, has the highest PoPS score in one of our GWAS loci, but was not the nearest gene and was therefore not prioritized (Supplementary Fig. 1).

Finally, we prioritized several genes with limited existing literature or functional studies pertaining to PD. Despite this, many of these genes have promising potential as drug targets for PD, *e.g.*, by repurposing existing drugs approved for other conditions (Supplementary Table 3). For example, inhibitors of exportin 1 (encoded by *XPO1*), a nuclear export protein crucial for maintaining cellular homeostasis, have been developed and approved for different cancer conditions^86^. *PIK3CA* encodes a lipid kinase essential for cell proliferation and survival and has an approved inhibitor for breast cancer^87^. We identified five additional genes (*EP300, MAP4K4, CAMK2D, NCOR1,* and *WDR43*) that may encode druggable proteins (Fig. 2). Among those, *MAP4K4* and *CAMK2D* were also prioritized by the L2G^20^ and Yu2024^21^ models (Fig. 1). Further, *EP300* is not only the nearest gene with the top local PoPS score, but there is also a non-synonymous variant (rs20551) in the credible set in high LD (R^2^ = 87%) with the lead variant (rs9611522).

**Figure 2.**
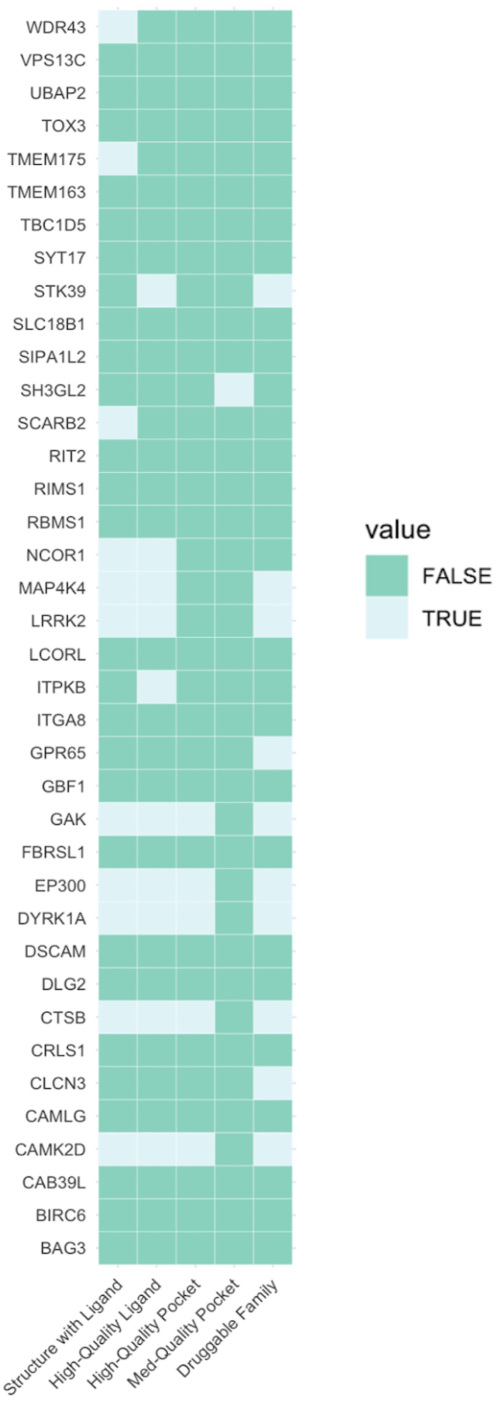
Small molecule target tractability assessment. Predicted tractability of the 38 prioritized genes that are not already targets of approved or investigational drugs. Data was extracted from the Open Targets platform using GraphQL API queries (https://platform.opentargets.org/) (see Methods). Various forms of evidence that suggest that a target may be tractable are shown on the x-axis, sorted from highest quality to lowest. Structure with Ligand: a Protein Data Bank co-crystal structure exists for the target and a small molecule. High-Quality Ligand: the target is bound by a ligand that 1) has a property forecast index ≤ 7, 2) binds ≤ 2 distinct protein domains and motifs identified by SMART (Simple Modular Architecture Research Tool), and 3) is derived from ≥ 2 distinct chemical scaffolds. High-Quality Pocket: the target has a DrugEBIlity score ≥ 0.7. Med-Quality Pocket: the target has a DrugEBIlity score 0–0.7. Druggable Family: the target was reported to be a member of the druggable genome in Finan *et al.* 2017^91^. Light green cells indicate that a given gene is supported by a given form of evidence, while dark green cells indicate an absence of such evidence. For more information on ongoing targeted drug trials for a selection of genes, see Supplementary Table 3.

**Figure 3.**
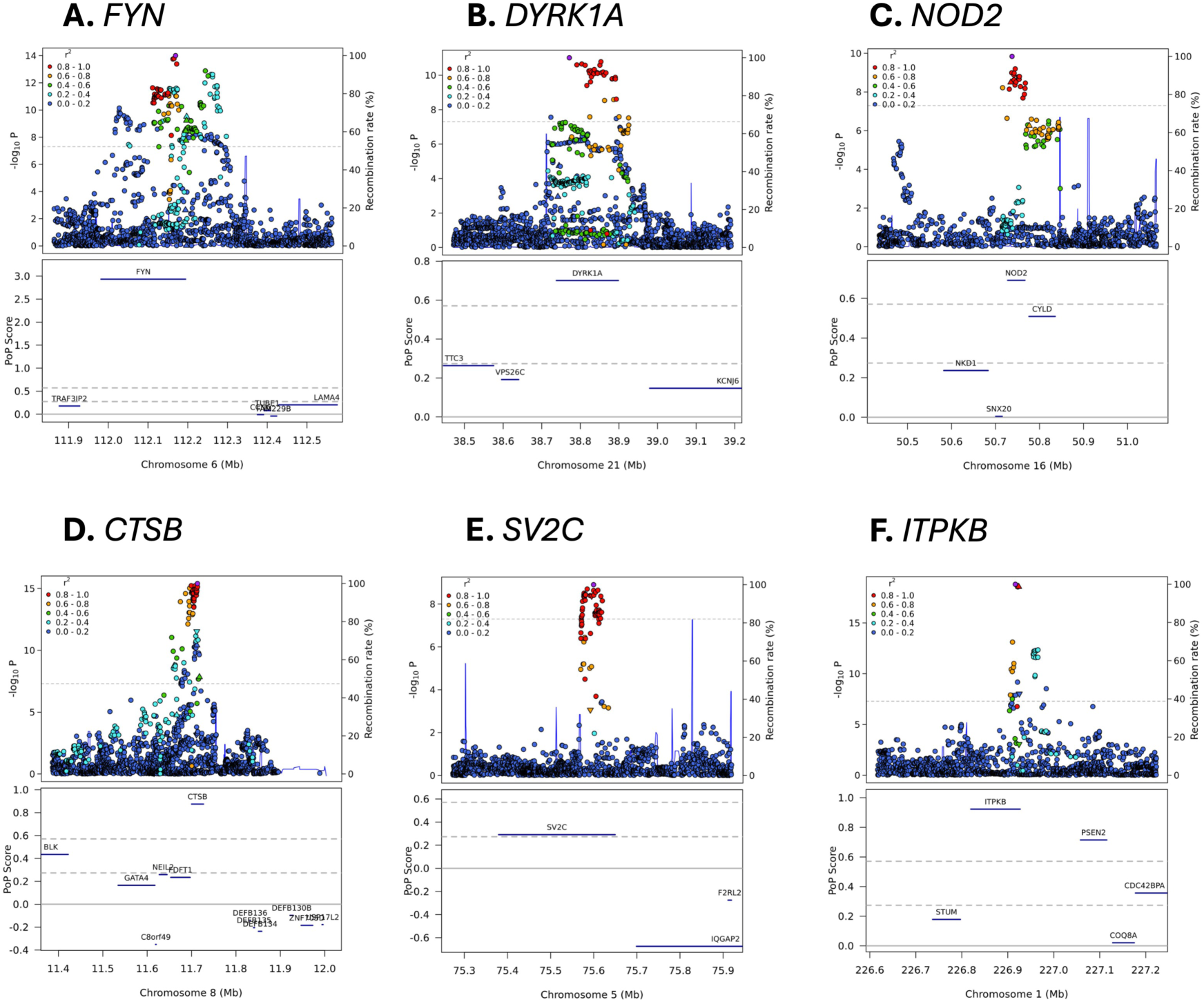
Variant-level associations and PoPS results for selected loci. The upper portion of each sub-plot is a LocusZoom plot. Each point represents a different genetic variant, the x-axis represents physical position on the listed chromosome, the left y-axis represents –log_10_-transformed P value, the right y-axis represents the recombination rate, colour represents linkage disequilibrium with the lead variant in the locus (as shown in the legend), and the horizontal dashed line represents the genome-wide significance P value threshold of 5×10^−8^. The lower portion of each figure is a PoPS plot. Genes are denoted as blue bars spanning from their transcription start site to their transcription stop site using the same x-axis as the LocusZoom plot, the y-axis represents the raw PoPS score, the dashed horizontal grey lines represent the top 10% and 1% of PoPS scores genome-wide, and the solid horizontal grey line represents a PoPS score of 0.

## Discussion

The field of targeted therapies driven by genetic discoveries for PD is rapidly evolving, with new therapies entering trials and ongoing studies bearing hope for more effective treatments. The goal of our study was to nominate high-confidence genes that warrant further follow-up investigations but bear promising potential as novel drug targets for PD. Following the established monogenic genes with ongoing trials already, *e.g., GBA1*, *LRRK2*, and *SNCA*, our prioritized genes may have the next-strongest links to PD and may represent our next-best opportunities for disease-modifying drug targets.

We prioritized 46 genes predicted to play a potentially important role in PD. In addition to prioritizing known and well-established PD genes, including *SNCA*, *LRRK2*, *GBA1*, *TMEM175* and *VPS13C,* we identified six genes (*FYN, DYRK1A, NOD2, CTSB, SV2C,* and *ITPKB*) emerged as promising targets for drug repurposing or novel therapeutic development. Our literature review found that each of these genes had a plausible mechanistic link to PD pathogenesis, paving the way for additional preclinical replication and validation experiments. We also nominated *XPO1, PIK3CA, EP300, MAP4K4, CAMK2D, NCOR1,* and *WDR43*. Despite limited prior research, these genes may be druggable and warrant further investigation. Lastly, we identified genes such as *RIT2, BAG3*, and *SCARB2,* with strong literature evidence supporting their involvement in neurodegenerative pathways linked to PD.

### The strength of PoPS compared to previous gene prioritization efforts

Several previous gene prioritization efforts have aimed to identify the causal genes in PD GWAS loci. For example, the PD GWAS browser is an online tool that displays data from a variety of resources for relevant PD risk loci, including GWAS statistics, eQTL, burden, expression, constraint, and literature data^88^. It serves as an exploratory scoring system that can be configured by users to assist them with the prioritization of genes located within PD GWAS loci. OmicSynth is a resource leveraging large-scale genetic and genomic data in a Mendelian randomization framework to identify therapeutic targets for neurodegenerative diseases^89^. The “locus-to-gene” (L2G) model is an XGBoost model trained on data from 445 curated gold-standard GWAS loci from 194 traits^20^. L2G integrates fine-mapping with functional genomics data, gene distance, and *in silico* functional predictions. The model has been applied to a large number of GWAS from a variety of traits, including the largest European-ancestry PD GWAS^13^. A similar but PD-specific XGBoost model was developed by Yu and colleagues (Yu2024)^21^. They trained their model on the largest European-ancestry PD GWAS^13^, using only seven well-established PD-associated genes as positive labels: *GBA1, LRRK2, SNCA, GCH1, MAPT, TMEM175,* and *VPS13C*. They used predictive features derived from gene expression and splicing in PD-relevant tissues such as brain, dopaminergic neuron subpopulations, and microglia.

The main advantage of the Yu2024 model over the L2G model is its PD-centred design. However, predictions made by the Yu2024 model appear to be poorly calibrated, possibly due to the small number of positive labels in its training set. For example, the largest European-ancestry PD GWAS^13^ identified a GWAS hit in strong linkage disequilibrium (LD, r^2^ = 97.9%) with a non-synonymous variant in the *FCGR2A* gene, strongly suggesting that this is the causal gene in the locus. Although conditional analyses revealed that this was the only independent signal in the locus, the Yu2024 model predicted that 19 genes in this locus had > 75% probability of being the causal gene with *FCGR2A* only ranking 18th.

All these previous efforts used a locus-by-locus approach, disregarding data from other GWAS loci and the remainder of the genome. In comparison, PoPS incorporates genome-wide information. The initial PoPS publication^19^ shows that PoPS outperformed other similarity-based and locus-based gene prioritization methods across numerous complex traits. By combining PoPS with nearest gene and non-synonymous credible set variant information, we prioritized a list of high-confidence genes as promising candidates for follow-up functional studies and potential drug development programs.

### Prioritization of genes with strong evidence supporting a mechanistic link to PD

Our work identified 46 genes that are very likely to be driving their signal in their respective GWAS loci. For a subset of these genes, there is also substantial literature supporting a mechanistic connection to PD. Some of these (*FYN*, *DYRK1A*, *NOD2*, *CTSB*, *SV2C*, and *ITPKB*) may serve as potential PD drug targets due to their involvement in pathways linked to neuroinflammation, autophagy, and neuronal degeneration. However, extensive further preclinical work is required to determine if and how these genes can be targeted and how this research can be translated into potential drug development. Another strategy for some of these genes is to evaluate how already existing drugs for other conditions may be repurposed for PD. Together, we hope these results stimulate interest in initiating drug development programs focused on these targets, all of which represent promising candidates for future PD therapies.

Approved FYN inhibitors could potentially be repurposed to reduce neuroinflammation and protein aggregation in PD patients^63–65^. Approved farnesyltransferase inhibitors could potentially be repurposed to enhance cathepsin activity, lysosomal function, and α-synuclein clearance^76,77^. *DYRK1A* inhibition may be a promising approach to investigate a potential reduction of α-synuclein aggregation and neuronal apoptosis. By developing MICU3 inhibitors, it may be possible to improve autophagy and reduce α-synuclein aggregation driven by elevated *ITPKB* activity. Although an approved drug already targets *NOD2*, it appears to have proinflammatory effects. By developing NOD2 inhibitors, it may be possible to decrease neuroinflammation and protect against dopaminergic neuron degeneration^79^. Additional research is also needed to determine the mechanism of action of investigational epilepsy drugs targeting SV2C, as they could potentially be repurposed for PD if confirmed as safe and effective activators.

### Prioritization of genes representing potentially druggable targets for PD

Furthermore, our approach identified a group of genes that has been relatively unexplored in PD. Our target tractability assessment indicated that some of these genes may be druggable targets, including *XPO1, PIK3CA, EP300, MAP4K4, CAMK2D, NCOR1,* and *WDR43*. Moreover, XPO1 and PIK3CA inhibitors, currently approved for the treatment of various cancer types, hold potential for repurposing as therapeutic agents for PD ^86,87^. We hope this first piece of evidence will stimulate further investigations and advocate for studies exploring the potential effects of overexpression and knockdown of these genes in cell and animal models for PD to better understand their potential mechanistic role. Such extensive preclinical work is crucial to facilitating and advancing drug development programs by pharmaceutical companies, and represents a critical step toward establishing these genes as viable therapeutic targets for PD.

### Limitations

Our study had several limitations. We were unable to assess genes on chromosome X because PoPS gene features are restricted to autosomes. Our approach failed to prioritize *GCH1,* a well-established PD risk gene^13,90^ implicated in dopamine synthesis in nigrostriatal cells. Although *GCH1* had the top PoPS value in its locus, the PIP-weighted centre of the credible set was slightly closer to the *WDHD1* gene. Previous machine learning gene prioritization efforts identified four genes with probability > 80% of being a causal PD gene that were not prioritized by our methods: *KCNS3*, *MBNL2*, *ASXL3*, and *SCAF11*^21^. However, there is only one genome-wide significant variant in the *KCNS3* locus (gnomAD non-Finnish European MAF = 9.7%) and all other variant P values are greater than 1×10^−5^, suggesting this may be a spurious association. *MBNL2*, *ASXL3*, and *SCAF11* were all the nearest gene in their loci, but did not have the top PoPS value.

Using GWAS data only from European and East Asian-ancestry could potentially restrict the generalizability of our findings across diverse genetic populations. The limited representation of these populations in GWAS highlights the need for larger studies in underrepresented groups to ensure equitable insights into PD genetics. Initiatives like the Global Parkinson’s Genetics Program (GP2) have been pivotal in addressing this gap, dedicating considerable efforts to advancing the study of underrepresented populations and fostering global collaborations in PD research.

### Conclusion

Gene prioritization efforts play a crucial role in nominating the underlying causal genes within GWAS risk loci, enabling the identification of potential novel drug targets. These efforts are essential for translating genetic discoveries into actionable therapies and have paved the way for personalized medicine approaches. Using PoPS and other methods, we prioritized a high-confidence list of 46 genes that are predicted to play an important role in PD pathogenesis and represent potential therapeutic targets. Prioritizing known and well-established PD genes, already targeted in clinical drug trials, strengthens the robustness and reliability of our approach. Newly prioritized genes may represent our next-best candidates for disease-modifying therapeutics. We hope our findings stimulate further investigations and preclinical work to facilitate drug development programs and potentially establish these genes as viable therapeutic targets for PD.

## Data Availability

All data produced in the present work are contained in the manuscript.

## Data availability statement

ChEMBL Database: https://www.ebi.ac.uk/chembl/

HRC reference release 1.1: https://ega-archive.org/datasets/EGAD00001002729

Gencode release 44: https://www.gencodegenes.org/human/release_44.html

OpenTargets platform: https://platform-docs.opentargets.org/

Details for accessing the EUR^13^ and EAS^16^ GWAS datasets can be found their original publications.

## Code availability statement

Custom code used in the presented study is stored at https://github.com/kheilbron/cojo_pipe and https://github.com/kheilbron/brett

Additional software and code:

COJO: https://yanglab.westlake.edu.cn/software/gcta/#COJO

MAGMA: https://cncr.nl/research/magma/

PLINK 1.9: https://www.cog-genomics.org/plink/

PoPS: https://github.com/FinucaneLab/pops

## Acknowledgements

This project was supported by the Global Parkinson’s Genetics Program (GP2). GP2 is funded by the Aligning Science Across Parkinson’s (ASAP) initiative and implemented by The Michael J. Fox Foundation for Parkinson’s Research (https://gp2.org). For a complete list of GP2 members see https://gp2.org. We would like to thank the research participants and employees of 23andMe, Inc. for making this work possible. We thank SURF (www.surf.nl) for the support in using the Snellius National Supercomputer. This work utilized the computational resources of the NIH HPC Biowulf cluster (https://hpc.nih.gov). This article has been posted on the medRxiv preprint server.

---

**Figure S1. Variant-level associations and PoPS results for the *MICU3* locus.** The upper portion of each sub-plot is a LocusZoom plot. Each point represents a different genetic variant, the x-axis represents physical position on the listed chromosome, the left y-axis represents –log_10_-transformed P value, the right y-axis represents the recombination rate, colour represents linkage disequilibrium with the lead variant in the locus (as shown in the legend), and the horizontal dashed line represents the genome-wide significance P value threshold of 5×10^−8^. The lower portion of each figure is a PoPS plot. Genes are denoted as blue bars spanning from their transcription start site to their transcription stop site using the same x-axis as the LocusZoom plot, the y-axis represents the raw PoPS score, the dashed horizontal grey lines represent the top 10% and 1% of PoPS scores genome-wide, and the solid horizontal grey line represents a PoPS score of 0.

