## Supplementary figures and images for "Prioritizing Parkinson’s disease risk genes in genome-wide association loci"

### Supplementary Figure

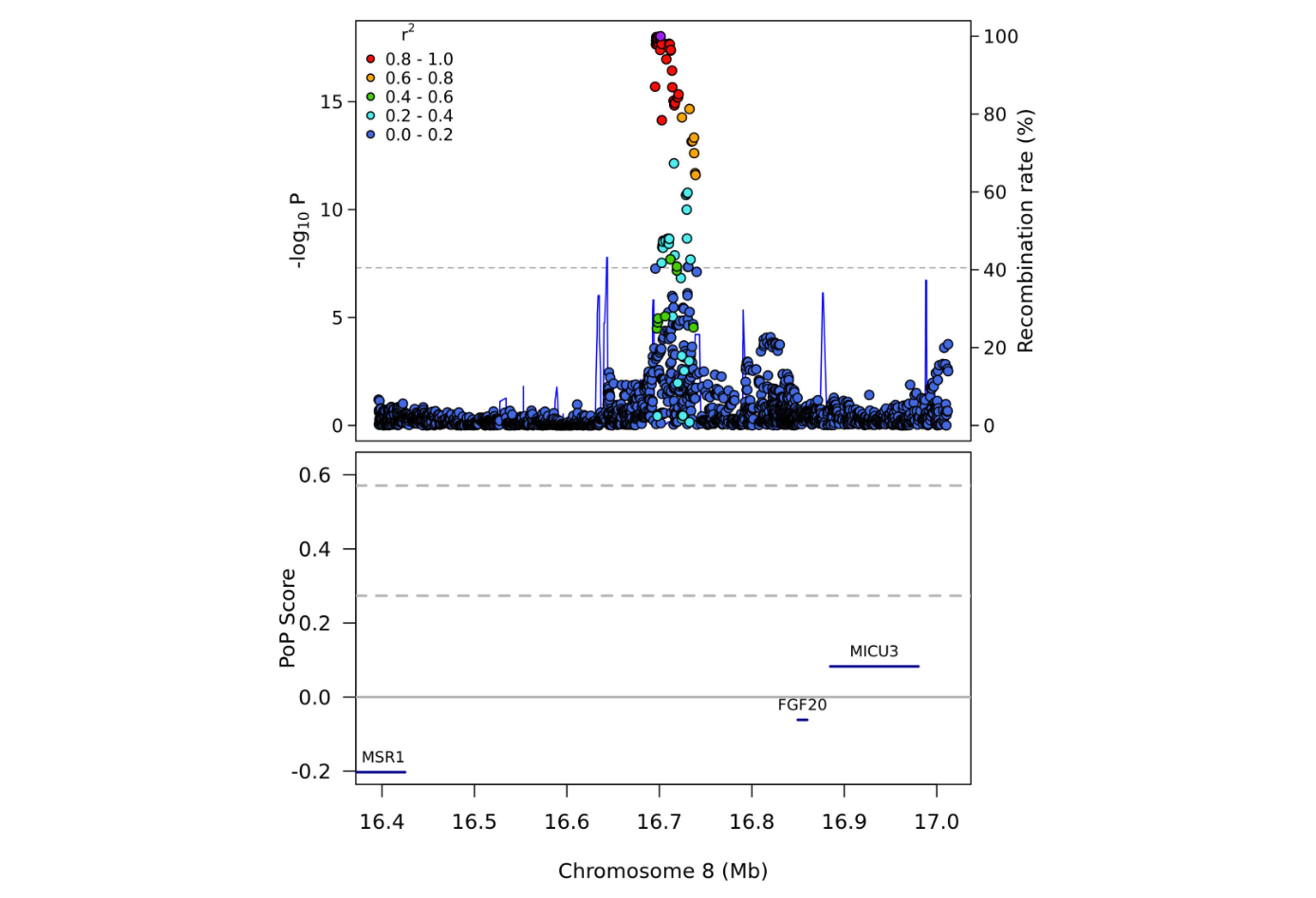
